# Usage Pattern and Associated Factors of Natural Mosquitoes Remedies in Endemic Communities of Borno State, Nigeria

**DOI:** 10.64898/2026.06.04.25342216

**Authors:** Raphael Kawep Njapdze, Idorenyin Bassey Ekerette

## Abstract

**Introduction:** Malaria, primarily transmitted by Anopheles mosquitoes, remains a major public health concern in Maiduguri, Borno State, Nigeria. While conventional control methods (e.g., ITNs) face challenges due to insecticide resistance and accessibility constraints, many communities rely on locally sourced natural products. This study aimed to assess the prevalence, usage patterns, and associated factors of these natural alternatives.

**Methods:** A cross-sectional survey was conducted across three purposefully selected communities in Maiduguri (Mairi, Furi, Lagos Street). A total of 450 household heads were interviewed using a structured questionnaire, collecting data on socio-demographics, specific natural products used, method of application, frequency, and perceived efficacy. Data were analyzed using descriptive statistics and binary logistic regression.

**Results:** Overall usage prevalence of natural products was high at 68.4%. The most common products identified were Neem (*Azadirachta indica*) extract (45.9%) and burnt Lemon Grass (*Cymbopogon citratus*) (31.2%). Usage pattern was predominantly indoor fumigation (burning), and over 70% of users prepared the products crudely at home. Logistic regression revealed that rural residence (Odds Ratio (OR): 2.1; p<0.01) and low education level (OR: 1.8; p<0.05) were significant independent predictors of higher natural product reliance.

**Conclusion:** Natural products constitute a widely adopted, community-driven vector control method in Borno State. The high prevalence and association with vulnerable populations suggest an urgent need to standardize the preparation and application of these products for potential integration into regional malaria control programs.

## INTRODUCTION

Mosquitoes are responsible for transmission of malaria (1, 2), Dengue fever (5), Zika virus (6), Yellow fever (7), Chikungunya (8), Filariasis (9), Japanese Encephalitis (10), West Nile virus (11). The World Health Organisation (WHO) estimated that half a million people are killed annually through infection of disease transmitted by mosquitoes (12). Malaria control is crucial for achieving United Nations Sustainable Development Goals (SDGs) 3 and 9 (13). Current control measures done to tackle the adverse effects of malaria in Nigeria include distribution of Insecticide-treated bed nets (ITNs), Indoor residual spraying (IRS), Artemisinin-based combination therapies (ACTs), Vaccination (RTSS) (14). Despite progress, challenges persist and these challenges range from resistance to insecticides and antimalarial drugs (15, 16), limited access to healthcare services, inadequate funding (17), Climate change and environmental factors (18). According to WHO, there were an estimated 236 million malarial cases (95% of global cases) and 590,935 malaria deaths (97% of global deaths) in African member states in 2022. Nigeria bears one of the highest diseases loads globally (19). In 2022, the WHO African region accounted for about 94% of global malaria cases (233 million), with Nigeria alone accounting for 27% of global cases (20, 21). Moreover, Nigeria accounted for 31% of all malaria deaths globally in 2022 (22). Borno State, located in the North-East geopolitical zone, presents a particularly challenging landscape for malaria control due to high endemicity, internal displacement, insecurity, and limited infrastructure, which restrict the consistent use of conventional tools like ITNs and IRS (23, 24). The escalating insecticide resistance in Anopheles populations, especially to pyrethroids used in ITNs, further necessitates the exploration of complementary control strategies (17, 25). Historically, communities have employed plant-based repellents and fumigants due to their local availability, low cost, and cultural acceptability (26, 27). Studies’ highlights the potential of various plant extracts and essential oils as mosquito repellents. Reports show that essential oils from Citronella (*Cymbopogon nardus*), Lemongrass (*Cymbopogon citratus*), and Catnip (*Nepeta cataria*) demonstrated high repellency against *Aedes aegypti* and *Anopheles gambiae* (28, 29, 30). Also plant extracts from Neem (*Azadirachta indica*), Tulsi (*Ocimum sanctum*), and Basil (*Ocimum basilicum*) have shown significant repellent activity against *Culex pipiens* (31, 32, 34, 33).

However, a significant knowledge gap exists concerning the actual usage patterns of these products in high-risk areas like Borno and the respiratory health risk of burning dried plant matter indoors (35). Previous studies have focused on laboratory efficacy, but little is known about who in the community is using them, which specific plant materials are favored, how they are prepared and applied, and what factors influence the decision to use them over conventional methods. The objective of this study was to assess the usage pattern and identify the socio-demographic factors associated with the use of natural repellents and adulticides against Anopheles mosquitoes in selected endemic communities of Maiduguri, Borno State, Nigeria.

## METHODS

### Study Area

This cross-sectional, community-based study was conducted in Borno State, Nigeria, between July and September 2024, corresponding to the peak malaria transmission season. Three Communities; one urban (Lagos Street), one semi-urban (Furi), and one rural (Mairi) (35), were purposefully selected to capture a diversity of socio-economic contexts and access to conventional control measures. This study was done following standard procedures put in place by the World Health Organization and there were no changes to the results after commencement.

**Fig 1:**
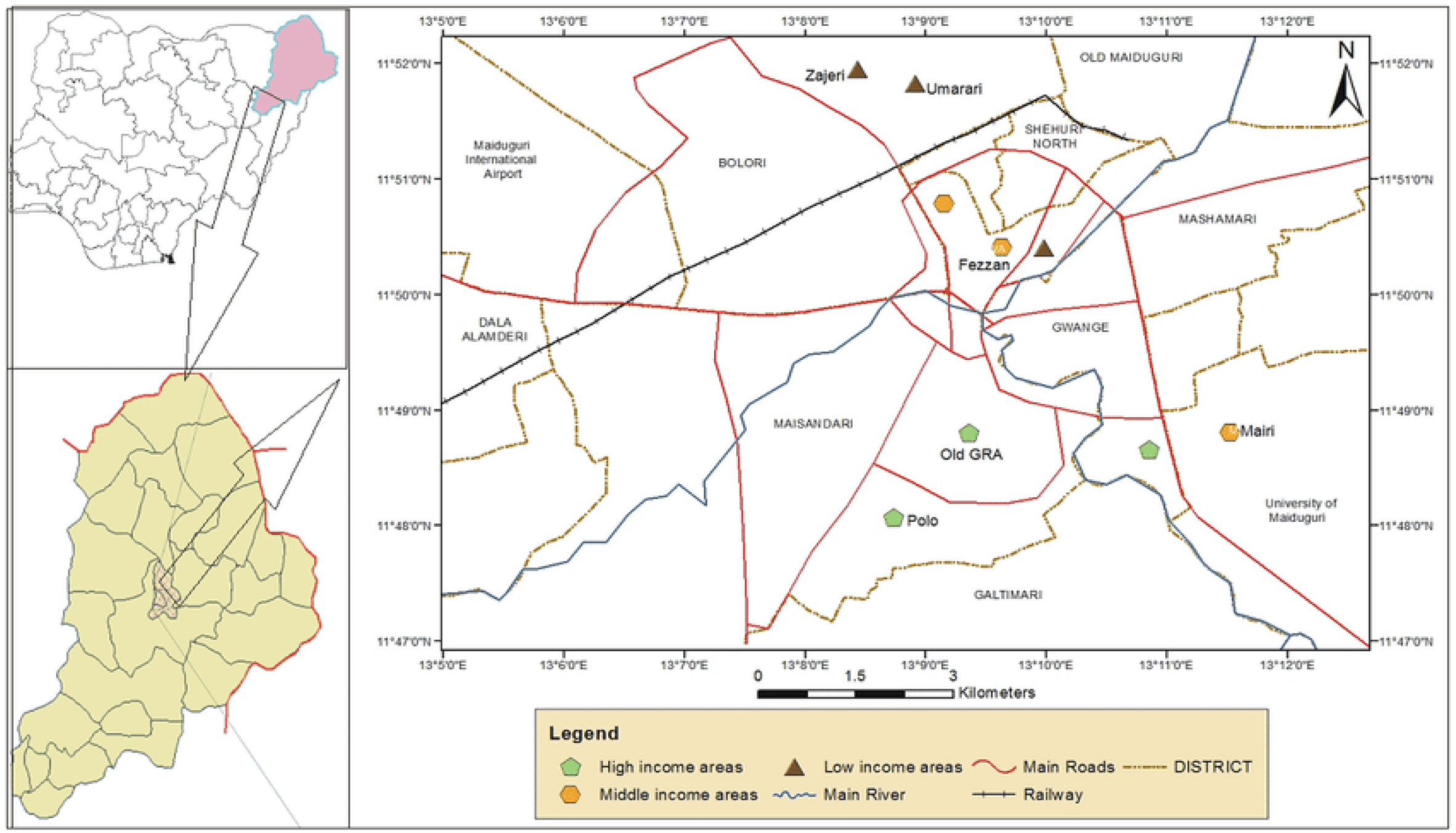
Map of Maiduguri with Selected Communities (Map of Study Area) *(Ref: Kelechi Friday Kwoncha)*

### Sampling and Participants

The target population included the head of households or a designated primary caregiver over 18 years of age. The minimum required sample size was calculated using the formula for estimating a single population proportion, assuming a 50% expected usage prevalence, a 95% confidence level, and a 5% margin of error, yielding a minimum of 384 respondents. Allowing for a 15% non-response rate, the final target sample size was set at 450 households. A multi-stage random sampling technique was employed. Within each selected LGA, wards and then settlements were randomly selected. Households were selected using systematic random sampling.

### Data Collection Instrument

A structured, pre-tested questionnaire translated into the dominant local language (Kanuri) was used for data collection. The questionnaire comprised four sections: (A) Socio-demographic data (age, gender, education, income, location); (B) Knowledge and practice of conventional control methods; (C) Usage of natural mosquito control products (types of plants/products, source, preparation method, frequency of use, and target application site); and (D) Perceived efficacy.

### Study Setting

Maiduguri lies between latitude 11°45□ N to 11°51□ N and longitude 13°2□ E to 13°9□ E. It’s located in the Ngadda Basin, a seasonal river that flows through Maiduguri. The rainy season starts and establishes in June and reaches its peak in August. Dry season start in October to May. Temperatures may go up to 48°C during dry periods and may drop to 15°C during the harmattan season. The mean annual relative humidity for dry and rainy seasons is 40% and 60% respectively while the mean annual evaporation rate is about 1600MM.

### Data Analysis

Completed questionnaires were checked for completeness and double-entered into Microsoft Excel and then exported to the Statistical Package for the Social Sciences (SPSS) version 26. Descriptive statistics (frequencies and percentages) were used to summarize socio-demographic data and usage prevalence. The Chi-square (*x*^2^) test was used to examine associations between socio-demographic variables (e.g., location, education) and the use/pattern of natural products. Variables found to be significant in the *x*^2^ analysis were entered into a Binary Logistic Regression model to identify independent predictors of natural product usage (P < 0.05).

## RESULTS

### Socio-Demographic Characteristics

Of the 450 questionnaires distributed, 437 were completed and included in the analysis (a response rate of 97.1%). The majority of respondents were female (61.8%) and household heads (75.3%). A significant proportion (42.1%) reported having only primary school education or less. Approximately 55% of the respondents lived in rural settings.

### Prevalence and Types of Natural Product Usage

The overall prevalence of natural product use against mosquitoes was high, with 299 respondents (68.4%) reporting current or past usage.

**Table 1:** Types of Natural Product Usage and Prevalence Amongst Users.

Types of Natural Product Usage and Prevalence Amongst Users
| Natural Product/Source | Primary Usage Form | Percentage of Users (N=299) |
| --- | --- | --- |
| *Azadirachta indica* (Neem) | Leaf/seed extract spray | 45.9% |
| *Cymbopogon citratus* (Lemon Grass) | Burnt dried leaves | 31.2% |
| *Ocimum gratissimum* (African Basil) | Topical application of crushed leaves | 12.7% |
| *Eucalyptus globulus* (Eucalyptus) | Burnt leaves/smoke | 6.1% |

Neem (*A. indica*) was the most frequently used product. The dominant mode of application was indoor fumigation via burning (48.5%) of dried plant material, followed by topical application (30.4%). Preparation was reported as “crude home preparation” by 70.6% of users, involving simple crushing or burning without standardization.

### Usage Patterns and Associated Factors

The analysis of usage patterns revealed clear variations across socio-demographic lines. The frequency of use was significantly associated with location. Daily usage was reported by 65% of rural respondents compared to 35% of urban respondents (*x*^2^ = 18.5, p < 0.001).

Binary Logistic Regression identified two significant independent predictors of natural product use.

**Table 3:** Natural Product Use in Rural and Urban Communities Involving High and Low Educated individuals.

| Predictor Variable |  | B | S.E. | Wald | p-value | Odds Ratio (OR) | 95% CI for OR |
| --- | --- | --- | --- | --- | --- | --- | --- |
| Rural vs Urban |  | 0.749 | 0.231 | 10.51 | 0.001 | 2.11 | (1.34, 3.32) |
| Low Ed vs High Ed |  | 0.587 | 0.252 | 5.43 | 0.020 | 1.80 | (1.10, 2.95) |

S.E = Standard error
B = Regression coefficient

The model indicates that respondents in rural areas were approximately 2.1 times more likely to rely on natural products than their urban counterparts. Similarly, individuals with lower education were 1.8 times more likely to use these alternatives.

## DISCUSSION

This study confirms that natural plant products are a major, community-driven component of vector control in Borno State, Nigeria, with over two-thirds of households reporting usage. The high reliance observed, particularly in rural settings, likely stems from factors critical to the region: low cost, accessibility, and high levels of displacement that often make obtaining conventional, subsidized ITNs and repellent sprays inconsistent. The prominence of Neem and Lemon Grass aligns with regional ethnobotanical studies that have demonstrated their high repellent and larvicidal properties in laboratory settings (27,32). However, our finding that the dominant usage pattern is crude indoor fumigation (burning) presents a complex public health challenge. While burning dried plant matter is a simple, cost-effective adulticide method, the resulting smoke can pose risks to respiratory health, particularly for children and the elderly (35). Furthermore, the lack of standardization means the active ingredient concentration is unknown, leading to variable and potentially sub-lethal efficacy, which may contribute to localized resistance development. The statistical association between rural residence and low education with higher reliance on these products reinforces the notion that these are methods of last resort for the most vulnerable populations.

This highlights a critical need for targeted public health interventions. Instead of dismissing these natural strategies, vector control programs should investigate methods to standardize the preparation of the most effective local botanicals (like Neem or Lemon Grass) into safer, regulated topical creams or sprays, or provide effective conventional alternatives consistently to these populations.

A primary limitation of this study is its cross-sectional design, which captures usage practice at a single point in time and relies on self-reported data, potentially leading to recall bias. Future longitudinal or intervention studies are warranted to assess the efficacy of these commonly used products in vivo and evaluate the impact of standardized natural repellent distribution on malaria incidence in these communities.

## CONCLUSION

The usage of natural repellents and adulticides is a widespread and significant community response to mosquito control challenges in Borno State, with high usage concentrated among rural and low-education populations. While the reliance on crude preparations poses public health risks, the high adoption rate signals an opportunity to integrate effective, standardized natural products into the regional vector management strategy.

## Data Availability

All data produced in the present study are available upon reasonable request to the authors

## ETHICAL CONSIDERATIONS

Ethical approval was obtained from the Department of Zoology University of Maiduguri, Maiduguri Borno State Nigeria. Verbal informed consent was secured from every participant before the interview commenced, and anonymity and confidentiality were maintained throughout the process.

